# Facing the COVID-19 epidemic in NYC: a stochastic agent-based model of various intervention strategies

**DOI:** 10.1101/2020.04.23.20076885

**Authors:** Nicolas Hoertel, Martin Blachier, Carlos Blanco, Mark Olfson, Marc Massetti, Frédéric Limosin, Henri Leleu

**Affiliations:** AP-HP.Centre, Paris University, Paris, France; INSERM U1266, Paris, France; Division of Biostatistics, Modeling and Health Economics, Public Health Expertise, Paris, France; National Institute on Drug Abuse, Bethesda, MD, 20892, USA; Columbia University/New York State Psychiatric Institute, 1051 Riverside Drive, Unit 69, New York, NY, 10032, USA

**Keywords:** COVID-19, SARS-CoV-2, mortality, ICU-bed occupancy, incidence, quarantine, lifting, screening, treatment, United States, New York

## Abstract

Global spread of coronavirus disease 2019 (COVID-19) has created an unprecedented infectious disease crisis worldwide. Despite uncertainties about COVID-19, model-based forecasting of competing mitigation measures on its course is urgently needed to inform mitigation policy. We used a stochastic agent-based microsimulation model of the COVID-19 epidemic in New York City and evaluated the potential impact of quarantine duration (from 4 to 16 weeks), quarantine lifting type (1-step lifting for all individuals versus a 2-step lifting according to age), post-quarantine screening, and use of a hypothetical effective treatment against COVID-19 on the disease’s cumulative incidence and mortality, and on ICU-bed occupancy. The source code of the model has been deposited in a public source code repository (GitHub®). The model calibrated well and variation of model parameter values had little impact on outcome estimates. While quarantine is efficient to contain the viral spread, it is unlikely to prevent a rebound of the epidemic once lifted. We projected that lifting quarantine in a single step for the full population would be unlikely to substantially lower the cumulative mortality, regardless of quarantine duration. By contrast, a two-step quarantine lifting according to age was associated with a substantially lower cumulative mortality and incidence, up to 71% and 23%, respectively, as well as lower ICU-bed occupancy. Although post-quarantine screening was associated with diminished epidemic rebound, this strategy may not prevent ICUs from being overcrowded. It may even become deleterious after a 2-step quarantine lifting according to age if the herd immunity effect does not had sufficient time to become established in the younger population when the quarantine is lifted for the older population. An effective treatment against COVID-19 would considerably reduce the consequences of the epidemic, even more so if ICU capacity is not exceeded.

## 1. Introduction

Global spread of coronavirus disease 2019 (COVID-19) has created an unprecedented infectious disease crisis worldwide. As of April 17, 2,074,529 confirmed cases and 139,378 deaths due to COVID-19, caused by the novel severe acute respiratory syndrome coronavirus 2 (SARS-CoV-2), had been reported worldwide *(1)*.

Even with uncertainties about COVID-19, such as the number of asymptomatic cases or the duration of the infectious period *(2)*, governments from endemic countries must put in place measures to minimize deaths and the economic impacts of viral spread. In this context, model-based predictions of the impact of different mitigation strategies can help policy makers make the right decisions in a timely way, based on the main objectives of mitigation. These objectives include minimizing mortality and morbidity, avoiding an epidemic peak to reduce the strain on healthcare services, flattening the epidemic curve to wait for antibody-based test for COVID-19, antiviral drug therapies and vaccine development, and mitigating the economic impact of the pandemic *(2)*. Such policy objectives may require different, sometimes competing measures *(3)*. Furthermore, the effect of mitigation strategies are likely to substantially differ by population characteristics, e.g., the proportions of elderly persons and people with chronic diseases who are at increased risk of severe infection *(4)*, healthcare system characteristics, e.g., intense care unit (ICU) bed availability, the stage of the epidemic, e.g., the number of people infected and immunized, the degree of population adherence to mitigation measures, the availability of human, economic and industrial resources, and possibly seasonality *(5)*. Therefore, model-based predictions of the potential impact of competing mitigation measures on the medical outcomes of the epidemic, taking into account local specificities of these parameters, can help support evidence-based policy decisions.

In the United States, New York City (NYC) is the most densely populated city and the hardest hit by the epidemic, with deaths rapidly increasing from 5 on February 29 to 4,168 on April 15 *(6)*. To face the epidemic, New York City has recently ordered all nonessential retailers and services to close or to keep all their workers home. New Yorkers are expected to largely stay home and wear a face covering when outside of home, and those who have symptoms or have tested positive for COVID-19 have been ordered to stay in quarantine. Many NYC hospitals are already overwhelmed while the epidemic is progressing in other regions of the United States. Although these measures are likely to slow down the viral spread and reduce substantially peak healthcare demand and deaths in the short term as shown by Ferguson et al. in their influential microsimulation study *(7, 8)*, the epidemic is predicted to rebound once these measures will be relaxed *(4, 7, 8)*. Therefore, projecting the impact of different possible mitigation measures that could be implemented at this stage, including quarantine extension, quarantine lifting for everyone or certain groups of people, post-quarantine screening and potential treatment progress on the medical outcomes of the epidemic is urgently needed *(2, 9)*.

Most prior simulation studies on SARS-CoV-2 infection *(9-16)*, with few exceptions *(7)*, have used SEIR (Susceptible, Exposed, Infectious, Recovered/Removed) compartmental models. Their results may be limited by the assumption that all individuals in the same compartment (e.g., infected people) share the same characteristics *(17)*. Although some SEIR studies took into account age- and location-specific risk of inter-individual contamination *(15, 16)*, stochastic agent-based microsimulation (ABM) models can improve estimation of the potential impact of mitigation measures by incorporating disease and individual characteristics, individual risks of infection and contagiousness, social interactions between individuals, and interactions over time *(18)*.

In this report, we performed a stochastic agent-based microsimulation model *(18-21)* of the COVID-19 epidemic in NYC and evaluated the potential impact of quarantine duration (from 4 to 16 weeks), type of quarantine lifting (1-step lifting for all individuals versus a 2-step lifting according to age), post-quarantine screening and use of a hypothetical effective treatment against COVID-19, on the disease’s cumulative incidence and mortality, and on ICU-bed occupancy. Because we are likely at least 1-year to 18 months away from substantial vaccine production *(2)*, we did not include this option in the present study. Advantages of ABM over other traditional modelling techniques include a flexible individual-based approach that can capture an emergent phenomenon with complex interactions between individuals in an heterogeneous population, and provide a natural description of a complex system *(18, 19)*. Because of several uncertainties that determine the risk of virus transmission, such as the number of asymptomatic cases and the duration of the infectious period *(2)*, the present analysis was based on a calibration process that accounts for several disease’s transmission parameters within the constraints defined by the contact matrix and known parameters of the disease.

## 2. Methods

Following previously described methods *(18-21)*, we performed a stochastic ABM model of the epidemic of COVID-19 in NYC. The model included 148 parameters summarized in **eTable 1**. Parameters on individual and disease characteristics (n=117) were mainly based on available data from prior studies and model calibration. Parameters related to social contacts were based on either prior studies (n=9) or assumptions when no data were available (n=22). The source code of the model has been deposited in a recognized public source code repository (GitHub®)

### 2.1. Individuals’ characteristics

The model was initialized with age (categorized by 5-year age groups) and household structures (proportions of singles, couples with children, couples without children, and single parents with children) observed in the NYC population, based on US Census Bureau data *(22)*. Households were distributed on a square grid that represents a geographical area. We used a single city grid since our study specifically focused on the COVID-19 epidemic in NYC. Based on data from the Department of Health of the New York State *(23)*, age and sex, all individuals were attributed a probability of having one or multiple diseases or conditions known to influence the risk of death from SARS-CoV-2, including smoking, hypertension, diabetes, coronary diseases, and chronic obstructive pulmonary disease *(24-26)*.

### 2.2. Social contacts

Social contacts were modeled to enable specific restrictions due to quarantine (e.g., school closure, public events’ cancellation), while conserving inevitable contacts such as with intrafamilial members or grocery shopping during the quarantine period. Given the complexity of modeling social contacts, we used a simplified set of contacts at both individual and household levels to model different types of social contacts experienced during the day *(27-30)*. These included close contacts for a prolonged duration with a small number of individuals, such as intrafamilial contacts, or people met at school or at work. They also included less frequent and less prolonged contacts with a finite set of individuals such as friends or extended family members. Finally, they included brief contacts with individuals in centralized locations such as grocery shopping, or in more remote locations such as when using public transport. For detailed parameters used to reproduce social contacts, please refer to *Supplemental text section*.

### 2.3. Disease’s characteristics

SARS-CoV-2 characteristics were mainly based on reports from the Centers for Disease Control and Prevention (CDC) *(6, 37)*, the European Centre for Disease Prevention and Control (ECDC) *(38)*, Santé Publique France reports *(39)*, and the London Imperial College reports *(40)*.

A key uncertainty about COVID-19 is the proportion of infected individuals that are not diagnosed. Studies from China *(26)* and Italy *(41)* suggest a high number of undiagnosed infections, ranging from 80% to 92% of all infections. Similarly, a study from France suggests that about 4 million people (representing about 6.0% of the French general population) were infected by the end of March 2020, contrasting with the 44,000 confirmed cases, suggesting a 1 in 100 diagnosis rate *(42)*. It was assumed that individuals with no or light symptoms (i.e., isolated minor symptoms of COVID-19 such as stomach aches, body aches, or nausea) were not diagnosed, except if they were traceable contacts (i.e., intrafamilial, work, school) of diagnosed patients, and that all individuals with mild, severe, or critical symptoms were diagnosed. To reflect these assumptions, among infected individuals, the probability of being asymptomatic or lightly symptomatic in the model was set at 100% in children aged less than 10 years, since very few children have been diagnosed with COVID-19 *(39)*; and was assumed to decrease linearly with age. The slope of this decrease was calibrated to show a cumulative incidence (diagnosed + undiagnosed) of 1 in 100 diagnosis rate.

The probability of severe and critical symptoms was based on hospitalization and mortality rates reported by the CDC *(37)*. Based on a prior study *(43)*, a 26% mortality rate was considered for patients in ICUs. Therefore, ICU occupancy was considered to be 4 times the mortality rate observed in ICUs. The prevalence of both severe and critical symptoms was exponentially distributed with age, with each disease or condition known to influence the risk of death from SARS-CoV-2 adding 5 years to the biological age. Delays between infection, symptom onset, hospital admission, ICU admission, death and recovery were based on prior reports *(17, 40, 44)* and are detailed in **eTable 1**. Delays were randomly assigned based on the Weibull distribution *(45)*.

The risk of infection during a contact with an infected individual (per min/m^2^ of contact) was calibrated to reproduce the SARS-CoV-2 epidemiological data from NYC until the 5^th^ of April *(46)*. A prior review *(47)* suggested that the median of the basic reproduction number (R_0_) of COVID-19 would be 2.79. This variable was included as an outcome in our model to examine whether the predicted value was in line with published reports, and thus evaluate the potential predictive value of the model. It was assumed that individuals with no symptoms or light symptoms had only a 10% risk of infecting other individuals to account for a probable lower viral load in these individuals. The risk of transmission was highest at the onset of the symptoms. To take into account the risk of transmission before developing symptoms *(17)*, it was assumed that infected individuals were contagious starting one day after infection, albeit with a contagiousness that decreases exponentially the further away from onset. An exponential function was chosen because it fitted well with the dynamics of viral replication, based on these assumptions. Individuals who recovered were considered as having acquired immunity against the virus and no longer at risk of infection. Based on prior work *(48)*, sensitivity of reverse transcription polymerase chain reactions (RT-PCR) to detect COVID-19 cases was assumed to be 71%.

Finally, the number of ICU beds needed over time was compared to the number of ICU beds available in NYC, estimated initially at 2.7 per 10,000 residents over the age of 15, i.e., 1,800 beds *(49)*. However, following healthcare systems’ reorganization and the opening of the Central Park’s Field Hospital, we assumed that ICU capacity in NYC has tripled from early April. Patients requiring ICU with no available beds were assumed to have 100% probability of dying.

### 2.4. Medical outcomes

Medical outcomes included cumulative incidence, cumulative mortality, and number of ICU beds needed.

### 2.5. Interventions

All diagnosed cases were assumed to have been quarantined. In the model, we also took into account efforts to track the contacts of diagnosed patients. Every intrafamilial, friend and family, work, and school contact of a diagnosed patient had in the previous days was systematically tested after an average delay of two days, representing the delay of the investigation. During this period, infected contacts could further spread the infection. People who met in grocery stores or in public transports were assumed to be untraceable. During quarantine, we considered that individuals had no contacts with other people, except with intrafamilial members and individuals at random in grocery stores and in streets.

We successively tested the following scenarios:

i. The natural course of the epidemic if no quarantine had been ordered.
ii. Different durations of quarantine: 4 weeks, 8 weeks, 12 weeks and 16 weeks.
iii. Two different types of quarantine lifting, i.e., for all individuals or a 2-step quarantine lifting according to age, i.e., a 3-week quarantine period for all individuals aged less than 60 years and an additional 8-week quarantine period for all individuals aged 60 years or more; a 70-year of age alternative cut-off instead of 60 years was also evaluated.
iv. Post-quarantine screening with RT-PCR tests of all symptomatic individuals and their contacts, and isolation of positive cases.
v. The use of a hypothetical effective treatment that would reduce the mortality by 90% of patients admitted to ICUs.

### 2.6. Statistical analyses

The stochastic agent-based microsimulation model (ABM) was run for 360 days from March 1, 2020 on 500,000 individuals. The results were extrapolated to 8.5 million individuals (i.e., the estimated NYC population). We examined whether the model had adequate calibration based on both two-sample Kolmogorov–Smirnov tests and visual comparison of the model-predicted and observed curves of the cumulative incidence of diagnosed cases and the cumulative mortality. We also examined whether the model-predicted value for the basic reproduction number (R_0_) of COVID-19, which was voluntarily considered unknown in our model, was in line with published reports.

We followed recent recommendations for the improvement of predictive mathematical models of the COVID-19 pandemic *(50)* and examined the robustness of our results by evaluating the impact on the estimated incidence and mortality of varying each model parameter value by +/-20%. These analyses were run for two-step quarantine lifting using a 70-year of age cut-off (i.e., <70y: 3-week-quarantine, ≥70y: 11-week quarantine) and for the differences in incidence and mortality between one-step 16-week quarantine for the full population and two-step quarantine lifting using a 70-year of age cut-off.

The model was performed using C++ and statistical analyses were conducted using SAS 9.4.

## 3. Results

### 3.1. Model calibration

**Figure 1** presents the results of the model calibration, supporting a good fit between observed and model-predicted cumulative incidence of diagnosed cases, cumulative mortality and age distribution of confirmed cases. Two-sample Kolmogorov–Smirnov tests comparing observed and model-predicted curves of incidence of diagnosed cases and mortality did not show significant differences [i.e., KSa=0.64 (p=0.81) and KSa=0.89 (p=0.89), respectively]. Finally, the R_0_ of COVID-19 predicted by our model was between 2.8 and 2.9 across the different scenarios tested, consistent with the findings of a review *(47)* of 12 studies suggesting that R_0_ estimates would range between 1.40 and 6.49, with a median of 2.79.

**Figure 1.**
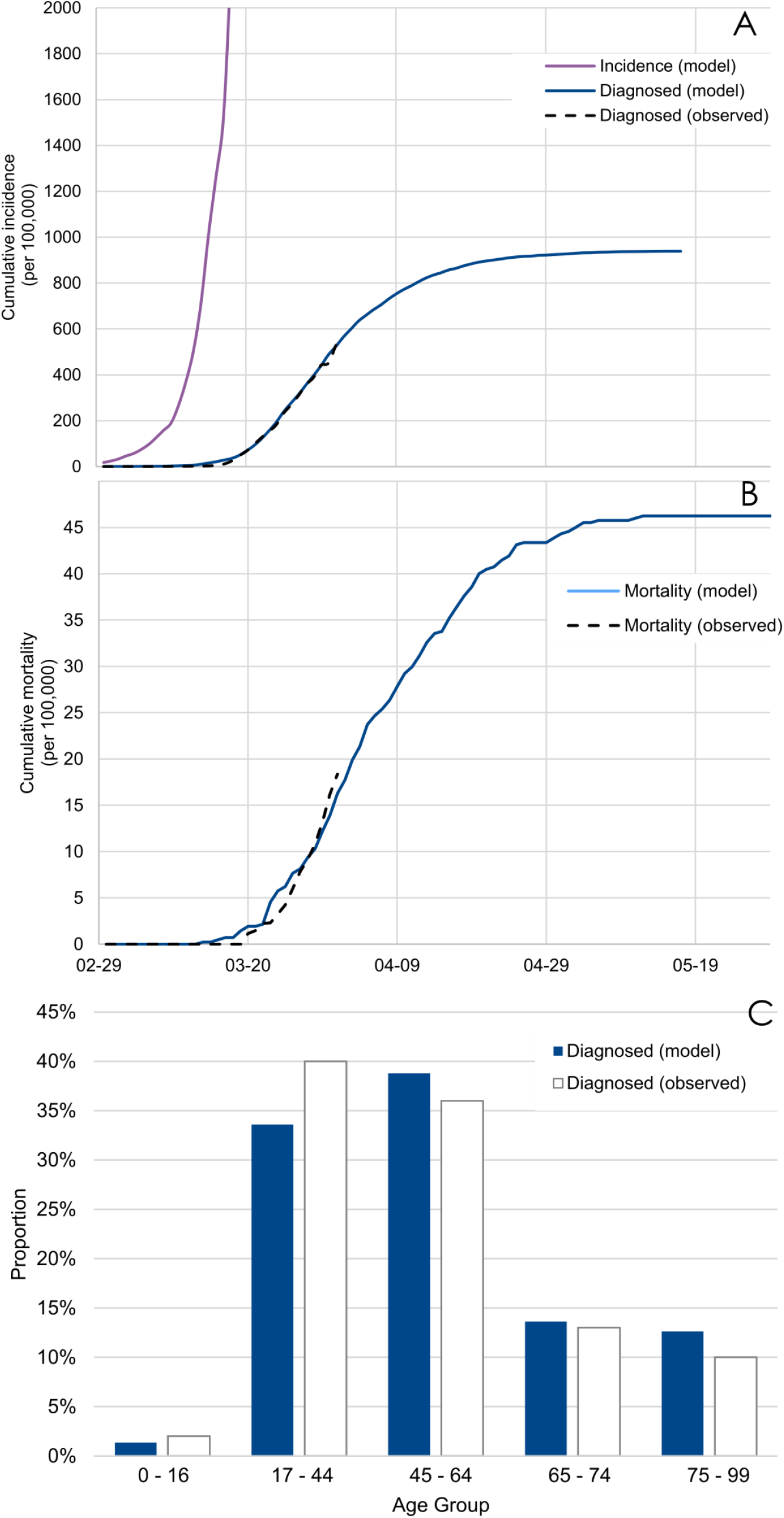
**Model-predicted and observed curves of the cumulative incidence of diagnosed cases (A) and the cumulative mortality (B), and age distribution of confirmed cases (C) in the epidemic of COVID-19 in New York City**.

### 3.2. Effect of quarantine duration

While quarantine was very efficient to contain the viral spread, we projected that it would be insufficient to prevent a second epidemic peak once lifted. Based on our model, the duration of quarantine was not associated with a reduced cumulative incidence or mortality, resulting in a similar, albeit delayed, overwhelming of ICUs (**Figure 2**).

**Figure 2.**
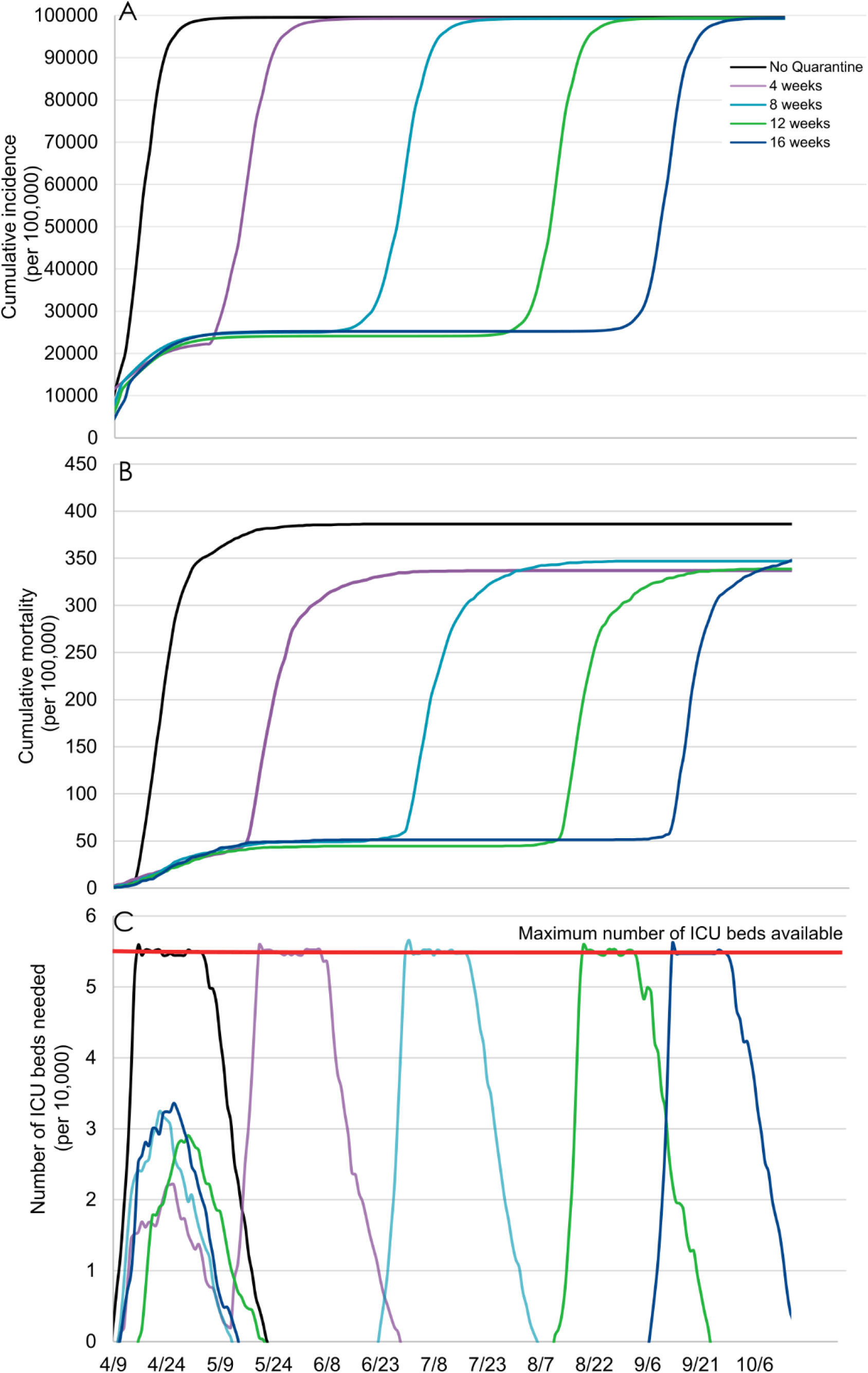
**Model-predicted cumulative incidence (A), cumulative mortality (B), and number of ICU beds needed (C) by quarantine duration, followed by a lifting for all individuals**.

### 3.3. Effect of one-step quarantine lifting for all individuals *versus* two-step quarantine lifting according to age

Our findings suggest that a 2-step quarantine lifting according to age, i.e., a 3-week quarantine for all individuals aged less than 70 years and an additional 8-week quarantine period for people aged 70 years or more, would lower the cumulative incidence by 23% and the cumulative mortality by 68%, compared to a 16-week quarantine followed by a lifting for all individuals (**Figure 3**). In addition, this strategy was not associated with overwhelming ICU bed capacity. The use of an alternative age cut-off of 60 years instead of 70 years resulted in an additional 3% decrease of the mortality, a substantial reduction of the number of ICU beds needed, and a similar incidence with a flattening of the cumulative incidence curve around 77% of the population.

**Figure 3.**
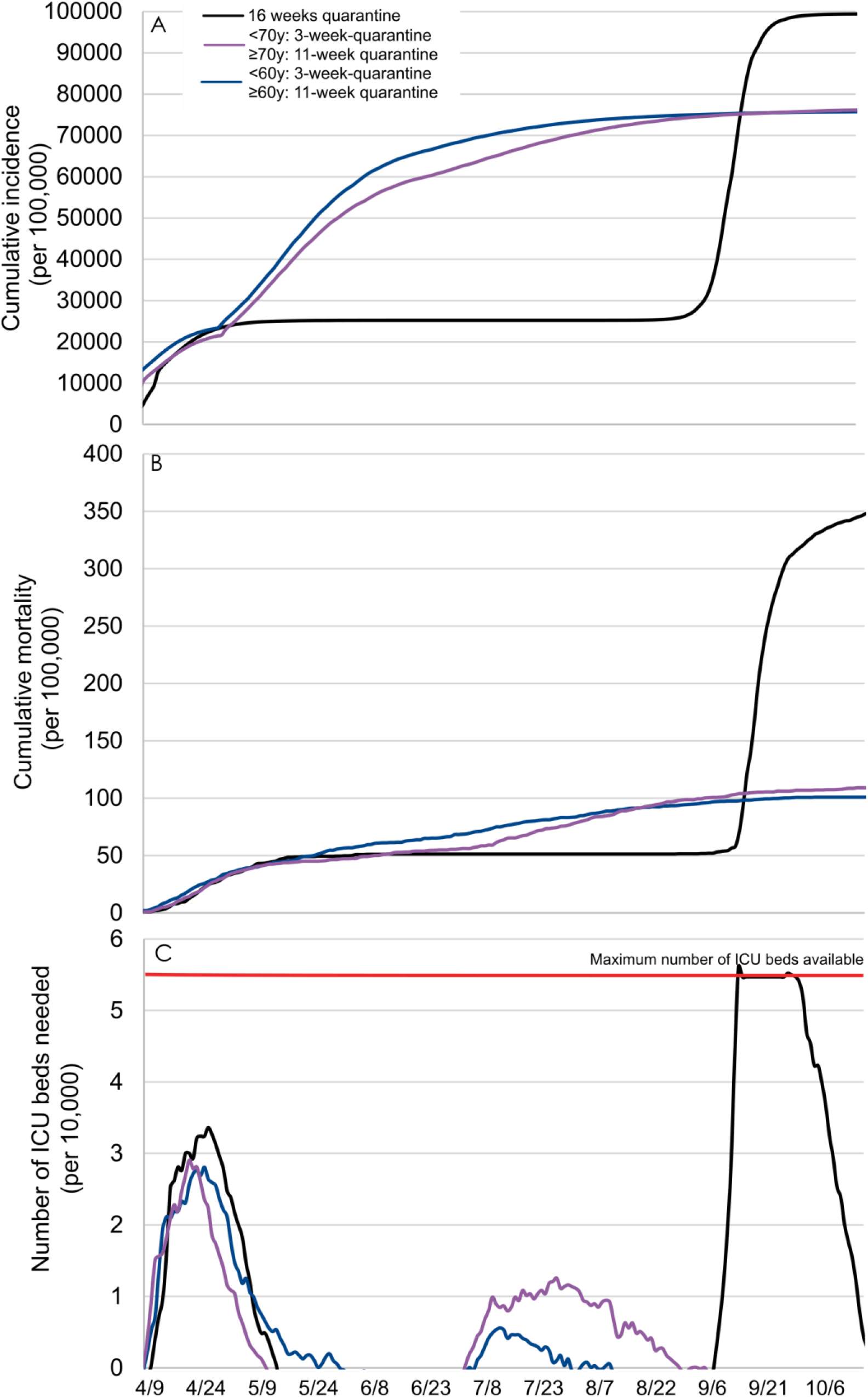
**Comparing model-predicted cumulative incidence (A), cumulative mortality (B), and number of ICU beds needed (C) between two-step quarantine lifting according to age, i.e**., **a 3-week quarantine for people aged less than 60/70 years and an additional 8-week quarantine for people aged 60/70 years or more, and one-step 16-week quarantine lifting for all individuals**.

### 3.4. Effect of the post-quarantine screening of all symptomatic individuals and their contacts with RT-PCR tests, and isolation of positive cases

We found that this strategy, after a 16-week quarantine followed by a lifting for all individuals, would be associated with a reduced mortality and cumulative incidence of 29% and 8%, respectively (**Figure 4**). However, this measure would be ineffective to prevent a second epidemic peak, likely to exceed available ICU beds. After a 2-step quarantine lifting according to age, this strategy was found to be useless or even potentially deleterious. Indeed, our model predicted that applying this strategy after a 3-week quarantine for all individuals aged less than 70 years and an additional 8-week quarantine period for people aged 70 years or more would result in a 18% increase in the mortality.

**Figure 4.**
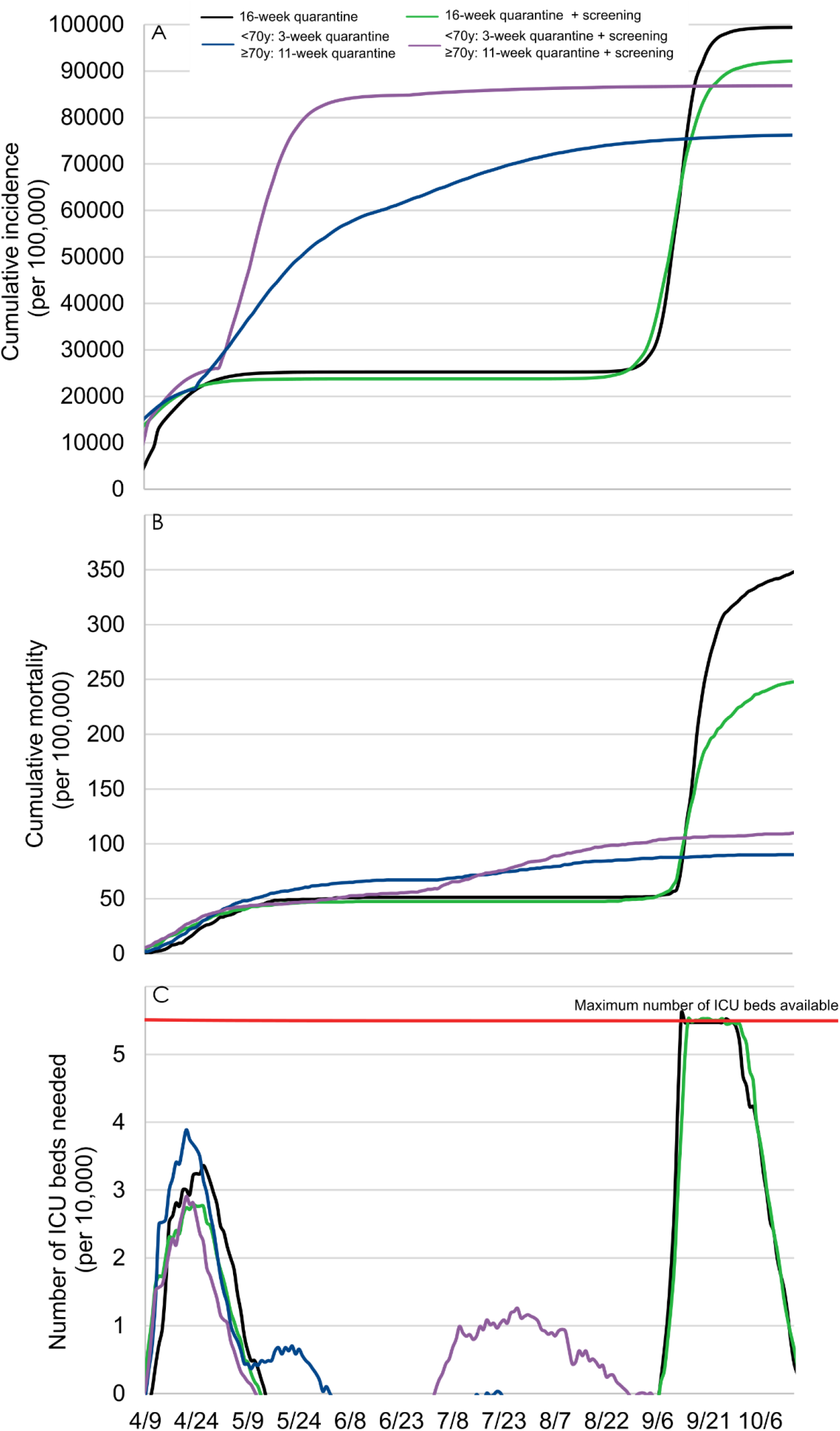
**Model-predicted cumulative incidence (A), cumulative mortality (B), and number of ICU beds needed (C) with an additional post-quarantine screening with RT-PCR tests of all symptomatic individuals**.

### 3.5. Effect of the use of a hypothetical treatment that would effectively treat 90% of patients with SARS-CoV-2 admitted to ICUs

The availability of an effective treatment for patients with SARS-CoV-2 admitted to ICUs would be highly beneficial to reduce the mortality after a 2-step quarantine lifting according to age using a 70-year of age cut-off, with an estimated mortality reduction of 88%. However, after a 16-week quarantine followed by a lifting for all individuals, the efficacy of this hypothetical treatment would be substantially reduced because of an overwhelming of ICUs, which would lead to a mortality reduction of only 29% due to the restricted proportion of patients likely to receive the treatment (**Figure 5**).

**Figure 5.**
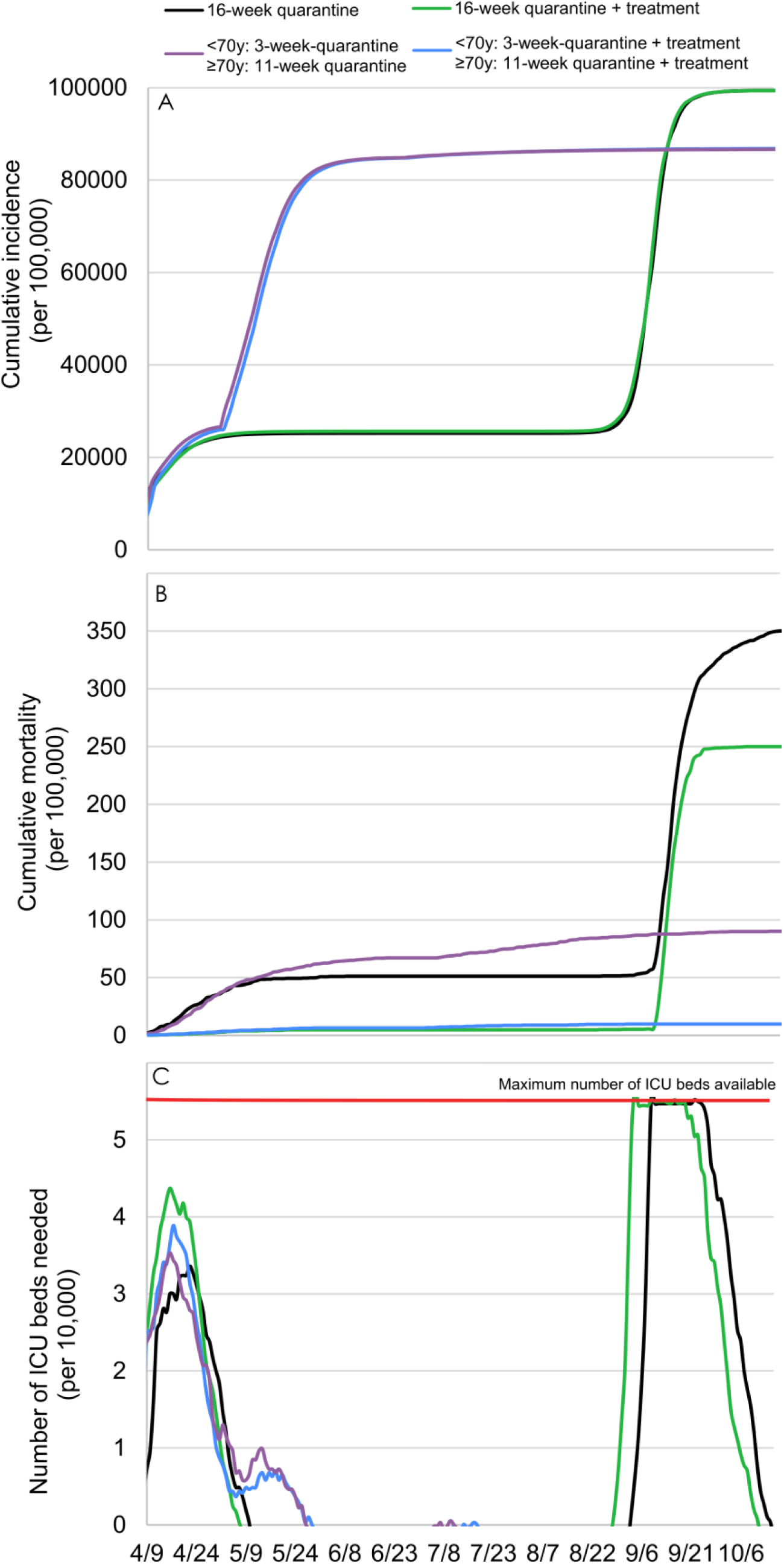
**Model-predicted cumulative incidence (A), cumulative mortality (B), and number of ICU beds needed (C) with the use of a hypothetical treatment that effectively treats 90% of patients with SARS-COV-2 in ICUs**.

### 3.6. Sensitivity analyses

When varying each model parameter value by +/-20%, we found that the estimated incidence and mortality would respectively change at most by 4,000 per 100,000 and 11 per 100,000 for a two-step quarantine lifting using a 70-year of age cut-off, and by 4% and 9% for the difference in incidence and mortality between a one-step lifting after a 16-week quarantine and a two-step quarantine lifting using a 70-year of age cut-off, suggesting the robustness of our results (**eFigures 1 to 4**). The only exception was the additional 3% decrease of the mortality when using an alternative age cut-off of 60 years instead of 70 years for a two-step quarantine lifting, which should thus be considered as marginal.

## 4. Discussion

To our knowledge, this is the first study to propose an agent-based microsimulation model of the epidemic of COVID-19 in New York City to predict the potential impact of quarantine duration (from 4 to 16 weeks), the type of quarantine lifting (1-step lifting for all individuals *versus* a 2-step lifting according to age), post-quarantine screening of all symptomatic individuals, and the use of a hypothetical effective treatment against COVID-19, on the disease’s cumulative incidence and mortality, and on the number of ICU beds needed. The variation of each model parameter value by ±20% had limited impact on outcome estimates (i.e., less than 4,000 per 100,000 for incidence and 11 per 100,000 for mortality), suggesting the robustness of our results. We projected that quarantine, while efficient to contain the viral spread, is unable to prevent a rebound of the epidemic once lifted, resulting in a similar, albeit delayed, overwhelming of ICUs. Based on our model, a 2-step quarantine lifting according to age, i.e., a 3-week quarantine period for younger individuals and an additional 8-week quarantine period for older people, would be associated with better outcomes, including a lower cumulative incidence, mortality and number of ICU beds needed, than a 16-week quarantine followed by a lifting for all individuals. An age cut-off of 60 years instead of 70 years did not substantially modify the mortality and incidence, but was associated with a reduced number of ICU beds needed. Although post-quarantine screening of all symptomatic individuals could bring some benefits after a quarantine lifting for all individuals, we found that this strategy may be ineffective after a 2-step quarantine lifting according to age, and potentially deleterious if herd immunity effect does not had sufficient time to be established in the younger population when the quarantine is lifted for the older population. Finally, an effective treatment against COVID-19 would considerably improve the consequences of the epidemic, even more so if ICU capacity is not exceeded.

Our findings reinforce that SARS-CoV-2 infection represents a major public health threat for NYC, as this infection may cause a very high number of deaths, estimated by our model at about 30,000 deaths in this city if no quarantine had been ordered and if ICUs had been overwhelmed. In line with prior work *(4, 7, 8)*, our findings suggest that quarantine, while an effective strategy to reduce the strain on healthcare systems by delaying the epidemic peak, is unable to prevent a rebound of the epidemic once lifted, regardless of its duration.

Importantly, based on our model, we found that a two-step quarantine lifting, i.e., a 3-week quarantine period for individuals aged less than 70 years and an additional 8-week quarantine period for people aged 70 years and over would be associated with better outcomes, including a lower cumulative incidence, mortality, and number of ICU beds needed, than a one-step quarantine lifting for all individuals, even when considering quarantine duration as long as 16 weeks. Furthermore, this strategy may potentially attenuate the severe economic and social consequences that a prolonged quarantine of the full population, including workers, would cause. As previously suggested *(51)*, these results could be explained by the herd immunity effect, i.e., the reduction of the infection as a result of the indirect protection observed in the unimmunized segment of the population in which a large proportion has been infected and therefore immunized *(51)*, as reflected by the predicted flattening of the cumulative incidence curve around 77% of the population with this strategy. Indeed, once the quarantine is lifted for younger healthy individuals in whom the risk of developing severe or critical symptoms or dying from SARS-CoV-2 is the lowest *(4)*, most of them would inevitably - in the absence of a vaccine - become infected and immunized, and could be adequately treated since ICUs are not expected to be overwhelmed, even during the peak incidence. Once most of them are immunized against COVID-19, the herd immunity effect is likely to prevent older adults from becoming infected when the quarantine would be secondarily lifted for them, i.e., 8 weeks later in our scenario. This strategy may prevent the risk of a second epidemic peak in this frailer population, and thus lessen subsequent mortality *(4)* and higher ICU-bed occupancy. Specifically, based on our model, the cumulative mortality in NYC would be expected to be about 9,350 deaths with this scenario, contrasting with the 29,750 deaths that would have been reached if the quarantine was lifted for all individuals after 16 weeks with no specific post-quarantine measures. When considering an alternative age cut-off of 60 years instead of 70 years, our model did not predict a substantial modification of mortality and incidence, but a substantial decrease in the number of ICU beds needed.

Although screening and isolation of symptomatic patients and of diagnosed patients’ contacts were associated with an attenuation of the epidemic course and a reduced mortality after a one-step 16-week quarantine lifting, this strategy may not prevent ICUs from being overwhelmed and would even be potentially deleterious after a 2-step quarantine lifting according to age. Specifically, in this scenario using a 70-year of age cut-off, this strategy would result in an 18% increase in mortality, probably because it delays the herd immunity effect in the younger population, leading a greater proportion of vulnerable older adults to be exposed to COVID-19. An extension of the quarantine period for the older people until the younger population is largely immunized would probably diminish this increased mortality. These findings may be explained by the assumption in the model and supported by prior studies *(41, 52)* that asymptomatic undiagnosed patients are responsible for a large hidden epidemic that exacerbates as soon as the confinement ends. Even when a much lower transmission rate was considered in the model for asymptomatic individuals, we found that disease progression persisted. However, the efficacy of this strategy is highly dependent on the assumption of the true diagnosis rate, which will be better estimated once results of large-scale studies using antibody-based tests for COVID-19 become available.

Our study has several limitations. First, the model was calibrated on the diagnosis and mortality rates available from the CDC *(6)*. However, we cannot exclude the possibility that these parameters are underestimates. Nevertheless, the observed differences across scenarios remained unchanged overall when considering a much higher and unlikely *(26, 41, 42)* diagnosis rate of 1 in 10, except for the efficacy of the post-quarantine screening on the course of the epidemic, which was much greater under this assumption, contributing to the robustness of our conclusions. Second, the contact matrix was approximated using multiple assumptions for each type of contact. However, we found that the model calibrated well, suggesting that although the assumptions made for unknown parameters, such as the frequency of meeting friends or the number of people met during shopping, can be criticized, the overall model, for which most parameter values were based on prior findings (**eTable 1**), may adequately predict the course of the COVID-19 epidemic in NYC. Third, we considered that infected people could develop immunity for at least several months following standard assumptions *(53)*. However, post-COVID-19 immunity length remains unknown *(54)*. Fourth, although the main differences observed were substantial and remained similar to a ±20% variation of each model parameter value, suggesting the generalizability of our results to other locales, future studies using this model and adjusting it to other city’s characteristics would be useful to verify this assumption. Finally, as with any simulation model, the results should be interpreted as estimates.

SARS-CoV-2 represents a major public health threat in NYC and worldwide. While quarantine is very efficient to contain the viral spread, we projected that it is insufficient to prevent a second epidemic peak once lifted. However, we found that a two-step quarantine lifting according to age was associated with a substantially lower cumulative mortality and incidence, up to 71% and 23%, respectively, as well as lower ICU-bed occupancy, than lifting quarantine in a single step for the full population. Although the post-quarantine screening was associated with a diminished epidemic rebound, our findings suggest that this strategy may not prevent ICUs from being overcrowded and may even become potentially deleterious after a 2-step quarantine lifting according to age if herd immunity effect does not had sufficient time to be established in the younger population when the quarantine is lifted for the older population. We hope that this model, the source code for which is publicly available, will be helpful to policy makers who face this devastating epidemic, and that the model will provide a useful framework to researchers to help inform future much-needed studies.

## Data Availability

The source code of the model has been deposited in a public source code repository (GitHub).

## Acknowledgments

We thank Pr Melanie Wall, Pr Yuanjia Wang and Mrs Marina Sánchez Rico for their helpful comments on early versions of this manuscript.

## SUPPLEMENTARY MATERIAL

**eTable 1.**
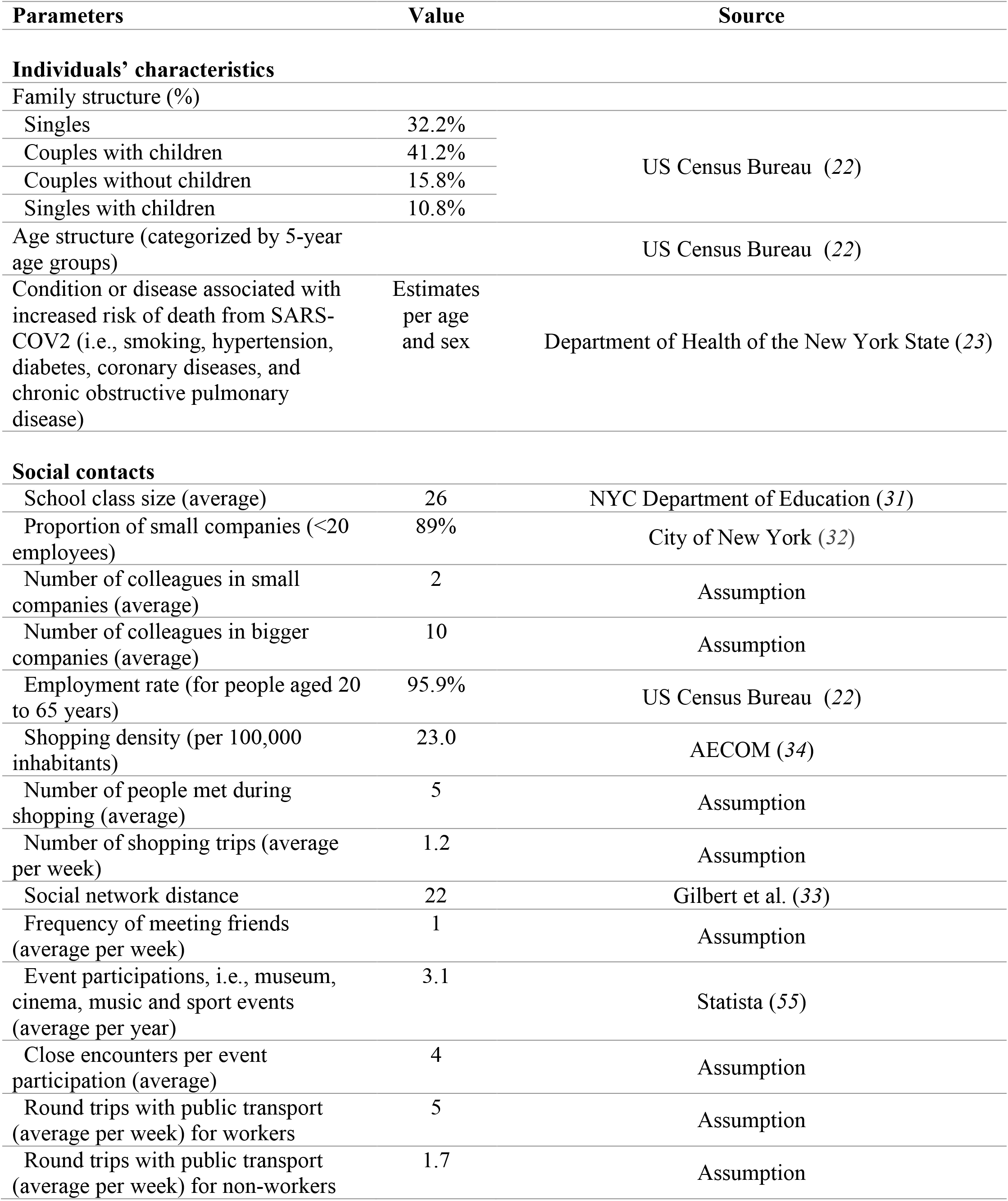
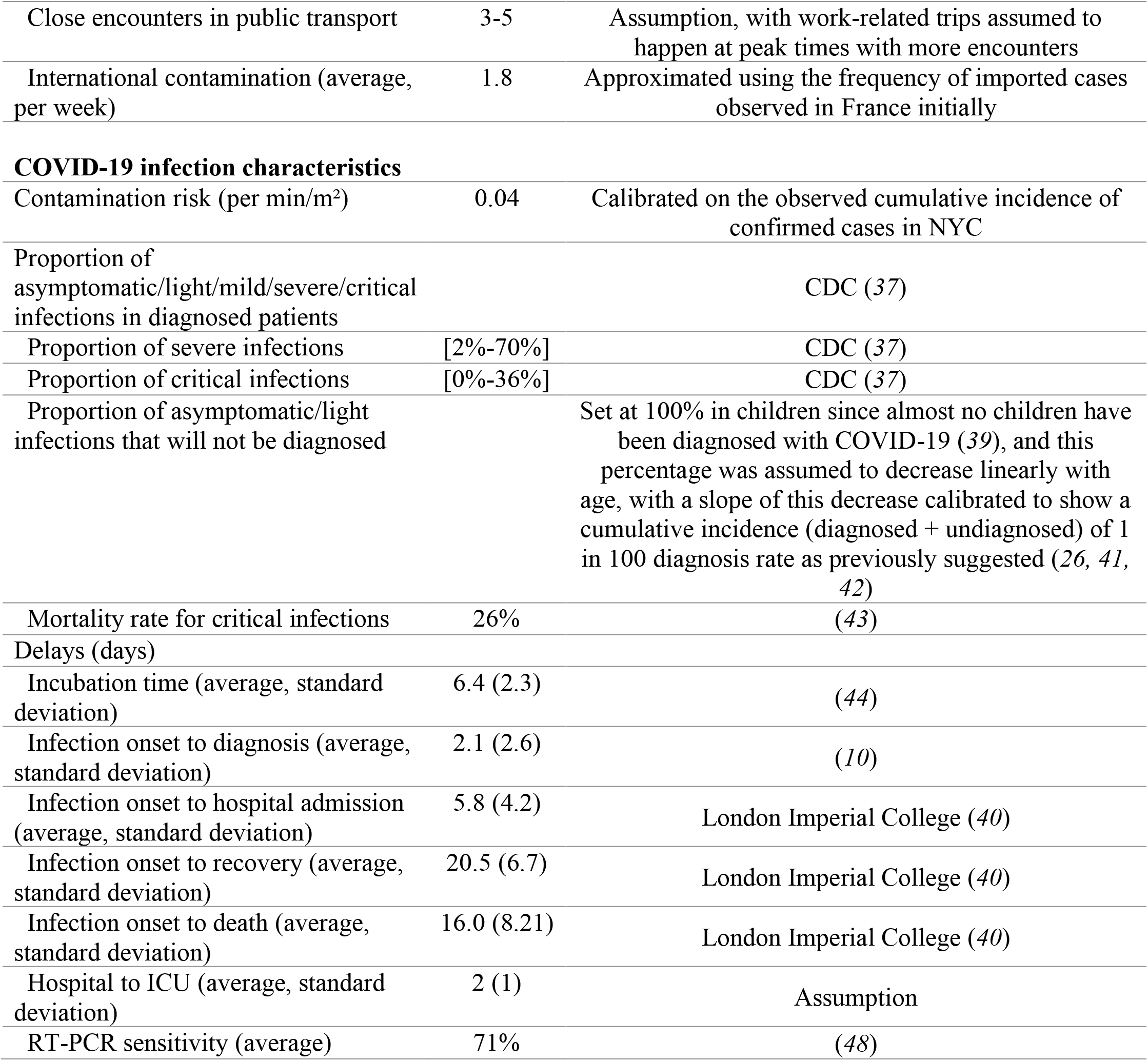
Summary of model parameters.

| Parameters | Value | Source |
| --- | --- | --- |
| **Individuals' characteristics** |  |  |
| Family structure (%) |  |  |
| Singles | 32.2% | US Census Bureau (22) |
| Couples with children | 41.2% |  |
| Couples without children | 15.8% |  |
| Singles with children | 10.8% |  |
| Age structure (categorized by 5-year age groups) |  | US Census Bureau (22) |
| Condition or disease associated with increased risk of death from SARS-COV2 (i.e., smoking, hypertension, diabetes, coronary diseases, and chronic obstructive pulmonary disease) | Estimates per age and sex | Department of Health of the New York State (23) |
| **Social contacts** |  |  |
| School class size (average) | 26 | NYC Department of Education (31) |
| Proportion of small companies (<20 employees) | 89% | City of New York (32) |
| Number of colleagues in small companies (average) | 2 | Assumption |
| Number of colleagues in bigger companies (average) | 10 | Assumption |
| Employment rate (for people aged 20 to 65 years) | 95.9% | US Census Bureau (22) |
| Shopping density (per 100,000 inhabitants) | 23.0 | AECOM (34) |
| Number of people met during shopping (average) | 5 | Assumption |
| Number of shopping trips (average per week) | 1.2 | Assumption |
| Social network distance | 22 | Gilbert et al. (33) |
| Frequency of meeting friends (average per week) | 1 | Assumption |
| Event participations, i.e., museum, cinema, music and sport events (average per year) | 3.1 | Statista (55) |
| Close encounters per event participation (average) | 4 | Assumption |
| Round trips with public transport (average per week) for workers | 5 | Assumption |
| Round trips with public transport (average per week) for non-workers | 1.7 | Assumption |
| Close encounters in public transport | 3-5 | Assumption, with work-related trips assumed to happen at peak times with more encounters |
| International contamination (average, per week) | 1.8 | Approximated using the frequency of imported cases observed in France initially |
| **COVID-19 infection characteristics** |  |  |
| Contamination risk (per min/m<sup>2</sup>) | 0.04 | Calibrated on the observed cumulative incidence of confirmed cases in NYC |
| Proportion of asymptomatic/light/mild/severe/critical infections in diagnosed patients |  | CDC (37) |
| Proportion of severe infections | [2%-70%] | CDC (37) |
| Proportion of critical infections | [0%-36%] | CDC (37) |
| Proportion of asymptomatic/light infections that will not be diagnosed |  | Set at 100% in children since almost no children have been diagnosed with COVID-19 (39), and this percentage was assumed to decrease linearly with age, with a slope of this decrease calibrated to show a cumulative incidence (diagnosed + undiagnosed) of 1 in 100 diagnosis rate as previously suggested (26, 41, 42) |
| Mortality rate for critical infections | 26% | (43) |
| Delays (days) |  |  |
| Incubation time (average, standard deviation) | 6.4 (2.3) | (44) |
| Infection onset to diagnosis (average, standard deviation) | 2.1 (2.6) | (10) |
| Infection onset to hospital admission (average, standard deviation) | 5.8 (4.2) | London Imperial College (40) |
| Infection onset to recovery (average, standard deviation) | 20.5 (6.7) | London Imperial College (40) |
| Infection onset to death (average, standard deviation) | 16.0 (8.21) | London Imperial College (40) |
| Hospital to ICU (average, standard deviation) | 2 (1) | Assumption |
| RT-PCR sensitivity (average) | 71% | (48) |

**eFigure 1.**
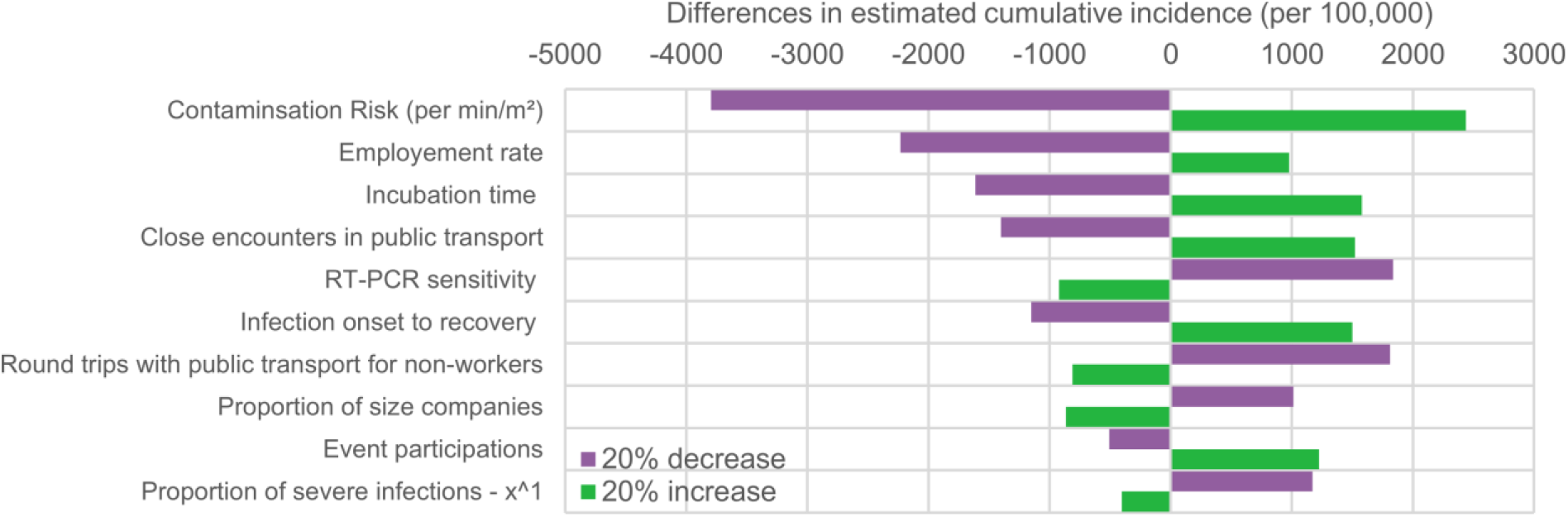
**Sensitivity analysis: impact of varying by +/-20% each model parameter value on the estimated cumulative incidence for a two-step quarantine lifting using a 70-year of age cut-off**. Only the 10 parameters having the highest impact are presented.

**eFigure 2.**
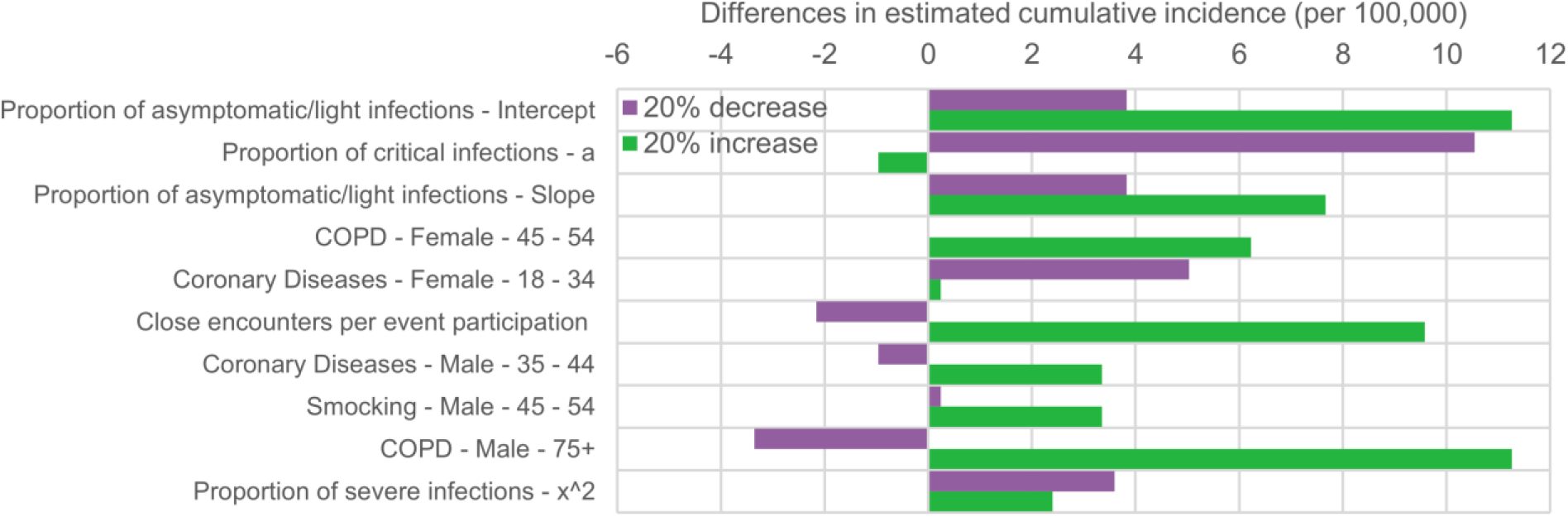
**Sensitivity analysis: impact of varying by +/-20% each model parameter value on the estimated cumulative mortality for a two-step quarantine lifting using a 70-year of age cut-off**. Only the 10 parameters having the highest impact are presented.

**eFigure 3.**
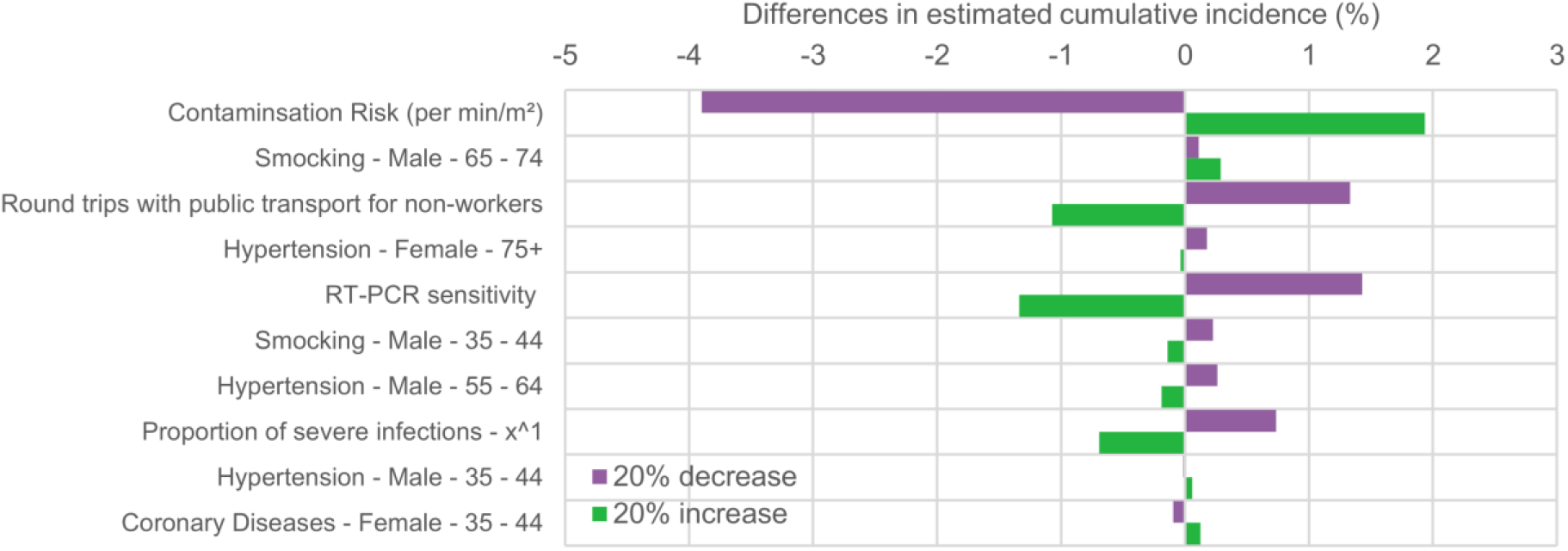
**Sensitivity analysis: impact of varying by +/-20% each model parameter value on the difference in cumulative incidence between a one-step 16-week quarantine and a two-step quarantine lifting using a 70-year of age cut-off**. Only the 10 parameters having the highest impact are presented.

**eFigure 4.**
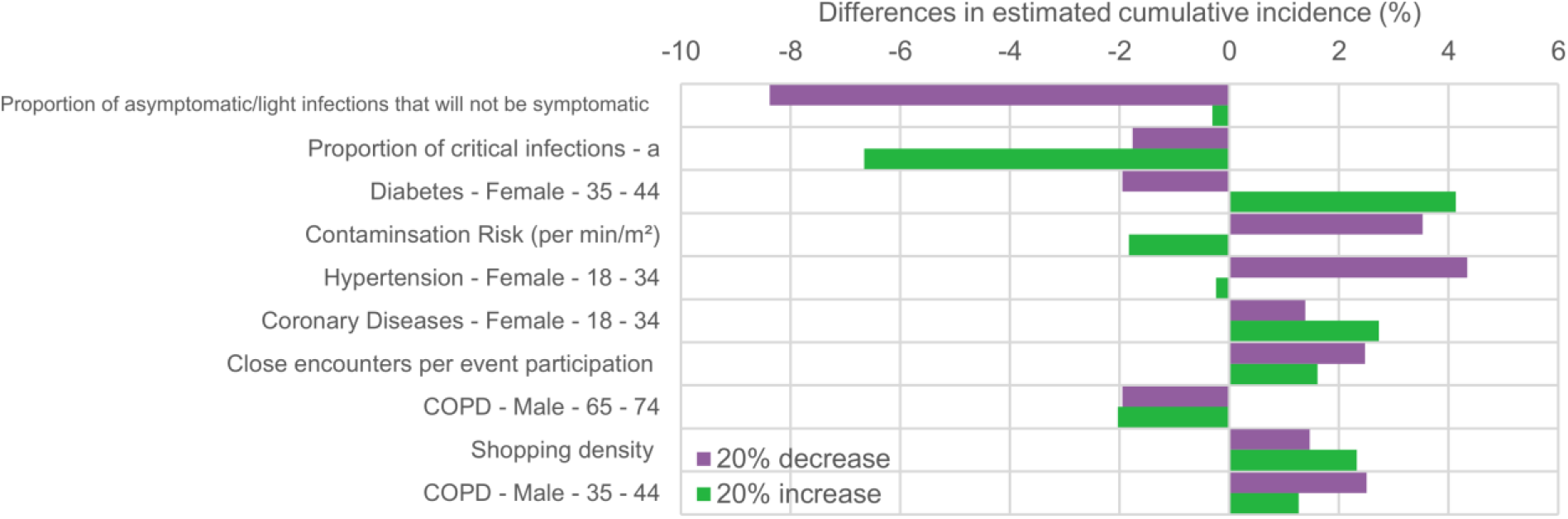
**Sensitivity analysis: impact of varying by +/-20% each model parameter value on the difference in cumulative mortality between a one-step 16-week quarantine and a two-step quarantine lifting using a 70-year of age cut-off**. Only the 10 parameters having the highest impact are presented.

## SUPPLEMENTAL TEXT SECTION

### Social contact model parameters

Contacts were defined by their average duration (in minutes), their average distance (in meters), their frequency, and the number of individuals involved. For intrafamilial contacts, it was assumed that their average duration was 6 hours per day at a 1-meter distance every day for all household members. For contacts at school, outside the quarantine period during which these contacts were considered null, average duration was 6 hours at an average 2-meter distance, 5 days a week, for all classmates. Classmates were identified as children of the same age living in a similar location to represent the geographic clustering of schools. The average class size in NYC was estimated at 26.1 *(31)*. For contacts at work, outside the quarantine period during which these contacts were considered null, average contact duration with colleagues was assumed to be 7.5 hours at a 2-meter distance, 5 times a week. Only employed individuals aged 20 to 65 years had work-related contacts. We distinguished between small companies with 20 or fewer employees and regular or large ones *(32)*. Individuals working in small companies had two colleagues on average, while employees of regular or large companies had an average of 10 colleagues. The number of colleagues was randomly drawn from a Poisson distribution. Work colleagues were identified at random within the city grid. For friends and family contacts, outside the quarantine period during which these contacts were considered null, it was assumed that the average duration was 180 minutes at a 1-meter distance, with one meeting a week on average. Outside the quarantine period, it was also considered that friend and family contacts occurred between households, for example, a couple with children could visit a friend’s or grandparent’s household.

Social networks were based on methods described by Gilbert et al. *(33)* with a distance of 22 (Poisson distributed) in order to incorporate key aspects of social networks, such as the different sizes of personal networks, high clustering, positive assortment of degree of connectivity, and low density. Individuals were considered to visit the closest grocery store from their location 1.2 times a week, and meet an average of five people (Poisson distributed). Grocery stores were uniformly distributed throughout the city grid and their density was estimated at 23 stores per 100,000 inhabitants *(34)*. Outside the quarantine period, contacts when going out of home were limited to cultural activities such as museum, sport, music or cinema events. It was assumed that contacts in restaurants or bars were captured through the friend and family contacts. The average number of times the family went out per year (Poisson distributed) was based on ticket sales’ from US statistics *(35)*. Attendance at any public event was associated with a duration of 120 minutes at a 2-meter distance with an average of 4 individuals (Poisson distributed) randomly identified in the city grid. We considered that all individuals used public transport 1.7 times a week for shopping or seeing family or friends. Workers were considered using public transports five times a week, twice a day (Poisson distributed). For public transport, a 30-minute average duration at a 1-meter distance from a mean number of 3 to 5 individuals (Poisson distributed) randomly identified in the city grid was assumed.

It was also considered that the first patients were individuals contaminated through international travel. Thus, individuals could become infected though international contacts over time at a rate based on the frequency of infected patients that were initially diagnosed in NYC *(6)*.

Finally, based on epidemiological data from South Korea *(36)*, it was assumed that the risk of transmission between individuals would be divided by four (representing an additional 1-meter distance for all contacts) if all individuals adhered to social distancing.

